# Associations between prenatal alcohol and tobacco exposure and cortical and subcortical brain measures in South African children: a pilot study

**DOI:** 10.1101/2022.06.07.22276078

**Authors:** Kristina A. Uban, Deborah Jonker, Kirsten A. Donald, Samantha J. Brooks, Stefanie C. Bodison, Eric Kan, Letitia Butler-Kruger, Annerine Roos, Babette Steigelmann, Brigitte Melly, Shana Adise, Andrew Marshall, Katherine L. Narr, Shantanu H. Joshi, Hein J. Odendaal, Elizabeth R. Sowell, Dan J. Stein

## Abstract

**Objective:** The aim of this pilot study was to assess associations of prenatal alcohol exposure (PAE), prenatal tobacco exposure (PTE), and their interaction and quantity on subsequent cortical and subcortical measures at age 6 years.

**Methods:** Mothers with varying levels of alcohol and tobacco exposure at different trimesters during pregnancy were approached when their children (born participating in the Safe Passage Study) were approximately 6 years old. 72 mothers agreed to participate, and 51 children completed brain magnetic resonance imaging (MRI). Brain regions of interest (ROIs) that were significantly associated prior to multiple comparison testing, were examined for associations related to exposure quantity, frequency, and timing (QFT), to explore how patterns of PAE and PTE influence brain outcomes in children. Linear regression was used to identify associations between PAE, PTE, and their interaction with cortical (n = 68 ROIs) and subcortical (n = 40 ROIs) measures.

**Results:** Prior to correction for multiple comparison testing, both PAE and PTE, as well as their interaction, were associated with a range of cortical and subcortical measures. However, none of these findings survived correction for multiple comparisons. Nevertheless, when exploring quantity of PAE, the total amount of standard drinks consumed during pregnancy and the average number of drinks per drinking day were positively associated with cortical volume in the right fusiform gyrus.

**Conclusion:** These trend results in this pilot study provide preliminary evidence that PAE impacts brain development in unique ways from PTE, and their interactive co-exposure is not a straight forward synergistic or additive effect on the brain.

## Introduction

Prenatal alcohol exposure (PAE) can produce enduring insults on the developing human brain, which can lead to challenges in function and health. Significant individual variability is observed in the range of these outcomes, due in part, to differing patterns of PAE and presence/absence of tobacco exposure (PTE) (Cook et al., 2015; McLachlan et al., 2017). Animal models clearly demonstrate alcohol as a teratogen, with variable PAE-effects on brain and cognitive outcomes that depend on quantity, frequency and timing (QFT) of PAE (Sulik, 1986; Sulik, 2008). While fetal alcohol spectrum disorder (FASD) is preventable, PAE is in the leading known cause of intellectual and developmental disability (May et al., 2013; Popova et al., 2016; Roozen et al., 2016; Lange et al., 2017), with an estimated 428 comorbidities (i.e., conduct disorder and receptive language disorder; Popova et al., 2016), making it a major global health concern.

There are several key gaps in FASD human literature. Firstly, most published studies examining the impact of PAE on the human brain have relied on retrospective designs for acquisition of maternal alcohol consumption patterns, introducing recall bias and error, and poor to no data collection capturing these patterns (quantity, frequency, timing; QFT). Secondly, much work has focused on clinical samples with heavy PAE versus unexposed participants, excluding mild and moderate exposure that occur within a given population. Thirdly, prenatal tobacco exposure (PTE) can also cause deleterious effects on brain and cognitive development (El Marroun et al., 2014; Wiebe et al., 2015; El Marroun et al., 2016), as shown in animal studies indicating interaction between PAE and PTE, (Bhattacharya et al., 2020); however, there are few human studies that examine this interaction (Odendaal et al., 2020). Fourthly, conducting magnetic resonance imaging (MRI) on young children aged 6 years old is challenging given introduction of motion artifacts in collected scans. Thus, this age range is limited in FASD human brain imaging literature for understanding early shifts in brain development.

The prevalence of FASD in the Cape region of South Africa is much higher than in many other national and international regions, with an estimate between 13.6 to 20.9% in a high-risk community sample of South African children in the first grade (May et al., 2013), compared to conservative estimates of 1-5% in the United States (May et al., 2018). This high prevalence reflects the history of South Africa, where there was systematic oppression of black farmworkers, including provision of alcohol in lieu of wages. Ongoing contributing factors to this risk may include prevailing socioeconomic disadvantages, psychological distress and depression, exposure to traumatic stressors, and intimate partner violence (Tomlinson et al., 2014; Stein et al., 2015; Koen et al., 2016; Donald et al., 2018).

A longitudinal community birth cohort, the Prenatal Alcohol, Sudden Infant Death Syndrome and Stillbirth (PASS) Network in this region, allows prospective investigation of the impact of a range of prenatal alcohol and potential co-occurring tobacco exposures. Working with PASS, we aimed to assess associations of PAE, PTE, and their interaction, with cortical and subcortical volumes at age 6, as well as associations between quantity of exposure and volumetrics. No other brain imaging study has examined the relationship between PAE, PTE and brain volumes in children aged 6. However, based on extant literature contrasting cortical volumes in FASD and neurotypical children (Nuñez et al., 2011), and in cross-sectional subcortical effects of PAE in older children versus neurotypical (Inkelis et al., 2020) it is hypothesised that significant interactions will occur in frontal and basal ganglia regions.

## Materials and methods

### Study design and participants

This research was a pilot sub-study embedded in the Safe Passage Study within the PASS Network, Western Cape, South Africa. This was a unique international community-based prospective birth cohort study investigating the role of PAE in the risk for sudden infant death syndrome (SIDS), stillbirth, Fetal Alcohol Syndrome (FAS) and FASD, funded by the National Institute of Child Health and Human Development (NICHD), the National Institute on Alcohol Abuse and Alcoholism (NIAAA), and the National Institute on Deafness and Other Communication Disorders (NIDCD).

As part of the Safe Passage Study of the PASS Network at Stellenbosch University, South Africa, 7,060 pregnant people were recruited from Bishop Lavis and Belhar residential areas between August 2007 and January 2015. These sites were selected on account of a historical reputation for having a high prevalence of PAE and SIDS and the need to include populations where the marked ethnic and socioeconomic disparities in SIDS remains understudied. Recruitment of pregnant women for the Safe Passage Study occurred between 6 weeks of gestation up to the day admitted for delivery. Methods and timelines are described in full elsewhere (Dukes et al., 2014).

The Safe Passage Study used a modified Timeline Follow-Back (TLFB) (Dukes et al., 2017), a validated method to capture alcohol exposure with a high level of detail, at the time around conception until the last day that the mother reported drinking. A validation study done by Himes and colleagues (2015) using a subset of Safe Passage Study women, indicated strong concordance between maternal reports using this approach and meconium biomarkers of alcohol exposure. The modified TLFB was administered at the recruitment interview (approximately 6 weeks GA), again at three different prenatal visits (20-24 weeks, 28-32 weeks, and 34+ weeks), and at one-month post-delivery. At the recruitment interview exposure information was collected for the time around conception (15 days before and after the last menstrual period) and for 30 days prior to the participant’s last reported drinking day. In subsequent interviews, if the participant reported consumption since her previous visit, the reference period consisted of the 30 days prior to the last drinking day. Peri-conception (2 weeks prior and 2 weeks following the last menstrual period) alcohol intake information was also collected. Detailed information was obtained to standardize and calculate the total grams of alcohol consumed on each drinking day or episode and detailed information regarding the type(s) of alcoholic beverage consumed was collected: whether the drink was frozen or included ice; number, and size of containers; the number of persons sharing; and interval of ingestion were collected for each drinking day.

For the purposes of this pilot study, 80 mothers with varying levels and timing of exposure in pregnancy and who had initially indicated that they would be willing to participate in future studies, were approached when their children (born as participants in the Safe Passage Study) were approximately 6 years old and invited to participate in the pilot study. All potential participants would necessarily have fulfilled the inclusion criteria of the Safe Passage Study, and these include:

1. Mother able to give consent;
2. Mother is at least 16 years of age at the time of consent;
3. Mother and child can speak Afrikaans and/or English. This criterion was not considered to unfairly exclude many research candidates: Afrikaans and English are the main spoken languages in the study region; thus, this criterion was included purely for practical reasons.

Additional exclusion criteria for children participating in the pilot study were:

1. History of traumatic brain injury with loss of consciousness exceeding 10 minutes;
2. Presence of a major medical or central nervous system disorder;
3. Maternal prenatal exposure to drugs (aside from tobacco and alcohol);
4. Implant (e.g., metal shunt) or medical condition that posed a risk during scanning.

Of the 80 randomly selected participants invited, 72 mothers agreed to participate. From the 72 participants, 51 children completed the scan.

### Calculating quantity, frequency, and timing of PAE and PTE

*The quantity* of PAE was measured using the total amount of standard drinks consumed during pregnancy and the average number of drinks per drinking day. The *frequency* of PAE was measured using the total number of days with binge consumption (more than 4 drinks per sitting). *Timing* of PAE identified the total amount of drinks consumed per trimester.

*The quantity* of PTE was measured using the total amount of cigarettes smoked throughout pregnancy. *The timing* of PTE was measured using the total amount of cigarettes smoked per trimester. There was no measure of frequency of PTE.

### Image acquisition

Scanning occurred at the Cape Universities Brain Imaging Center (CUBIC) located at Tygerberg Hospital, Cape Town. Whole-brain T1-weighted images were acquired for each participant using a 3-Tesla Siemens Allegra scanner. Scan parameters were as follows: repetition time (TR) = 2530 ms; echo time (TE) = 6.53 ms; flip angle = 7°; field of view = 224×168 mm^2^; voxel = 1×1×1 mm. Children were familiarized with the MRI environment during a session in a mock scanner on the same day prior to the actual scanning session.

### Image processing

FreeSurfer’s v5.3 recon-all pipeline was used to perform volumetric segmentation. This pipeline involve motion correction and averaging (Reuter et al., 2010) of multiple volumetric T1 weighted images (when more than one was available), removal of non-brain tissue using a hybrid watershed/surface deformation procedure (Ségonne et al., 2007), automated Talairach transformation, segmentation of the subcortical white matter and deep gray matter volumetric structures (including hippocampus, amygdala, caudate, putamen, ventricles) (Fischl et al., 2004a; Fischl & Dale, 2000), intensity normalization (Sled et al., 1998), tessellation of the gray matter to white matter boundary, automated topology correction (Fischl et al., 2002; Ségonne et al., 2007), and surface deformation following intensity gradients to optimally place the gray/white and gray/cerebrospinal fluid borders at the location where the greatest shift in intensity defines the transition to the other tissue class (Dale & Sereno, 1993; Dale et al., 1999; Fischl & Dale, 2000). Once the cortical models are complete, a number of deformable procedures can be performed for further data processing and analysis including surface inflation (Fischl et al., 1999a), registration to a spherical atlas which is based on individual cortical folding patterns to match cortical geometry across subjects (Fischl et al., 1999b), parcellation of the cerebral cortex into units with respect to gyral and sulcal structure (Fischl et al., 2004b; Desikan et al., 2006), and creation of a variety of surface based data including maps of curvature and sulcal depth. Whole brain metrics for cortical and subcortical volume Regions of Interest (ROI) were extracted using the Desikan-Kiliany atlas in tabular format. FreeSurfer’s subcortical pipeline segments each voxel into one of approximately 40 ROIs. These include the following bilateral structures: cerebral white matter, cerebral cortex, lateral ventricles, inferior lateral ventricles, cerebellum white matter, cerebellum cortex, thalamus, caudate, putamen, pallidum, hippocampus, amygdala, and accumbens area as well as midline and specialized labels such as: lesions, vessels, 3^rd^ ventricle, 4^th^ventricle, brain stem, and cerebrospinal fluid. These ROIs include the caudate of the left and right hemisphere, putamen of the left hemisphere, anterior cincular cortex and the cerebellum of the right hemisphere. The Deskian-Kiliany atlas was used to parcellate the cortical surface into 68 ROIs. Cortical volume, thickness and surface area metrics were calculated for each ROI.

### Statistical analyses

All statistical analyses were performed in CRAN R v4.1. Linear regression was run in R with the lm() package to identify associations between subcortical volume and prenatal exposure (PAE, PTE) as well as the PAE × PTE interaction. The regression model included age (months) and biological sex, and intracranial volume (ICV) as factors. The main effects and interactions were considered statistically significant at p < 0.05. The Benjamini-Hochberg false-discovery rate (FDR) correction (q < 0.05) was applied within each metric group.

## Results

Due to missing PTE information, four children with PAE were removed from the analysis. The final sample for analysis included 30 children with PAE and 17 control children without PAE (see Table 1). There was no significant difference in sex, household income and birth weight of the children between the PAE and control groups. However maternal smoking status was significantly different between the groups, where the ratio of smokers to non-smokers was higher in the PAE group (n = 21) compared to the control group (n = 7), p = 0.05. On average, the total number of alcoholic drinks consumed were highest in the first and second trimester and reduced in the third trimester. In contrast, tobacco use was consistent throughout pregnancy for both PAE and non-PAE groups, and at consistently higher doses of tobacco quantity among the PAE group relative to non-PAE group across all three trimesters (p’s < 0.05). A description of exposure information can be found in Table 2.

**Table 1.** Descriptive characteristics of the participants

| Variables | Control group (N=17) | PAE group (N=30) | *p*-value |
| --- | --- | --- | --- |
| **Sex** |  |  | 0.679 |
| Male | 9 (52.9%) | 14 (46.7%) |  |
| Female | 8 (47.1%) | 16 (53.3%) |  |
| **Household income (ZAR)** |  |  | 0.152 |
| N-Miss | 1 | 4 |  |
| Mean (SD) | 450.347 (311.997) | 653.399 (498.194) |  |
| Range | 71.429 - 1000.000 | 71.429 - 2000.000 |  |
| **Birth weight** |  |  | 0.724 |
| N-Miss | 0 | 2 |  |
| Mean (SD) | 3155.412 (503.669) | 3100.357 (503.899) |  |
| Range | 2140.000 - 3880.000 | 1800.000 - 3720.000 |  |

Note: Data are presented as $n/N$ (%) or mean (Standard Deviation) and range (lowest to highest values). Missing data: household income ( $n = 5$ ); birth weight ( $n = 2$ ).

**Table 2.**
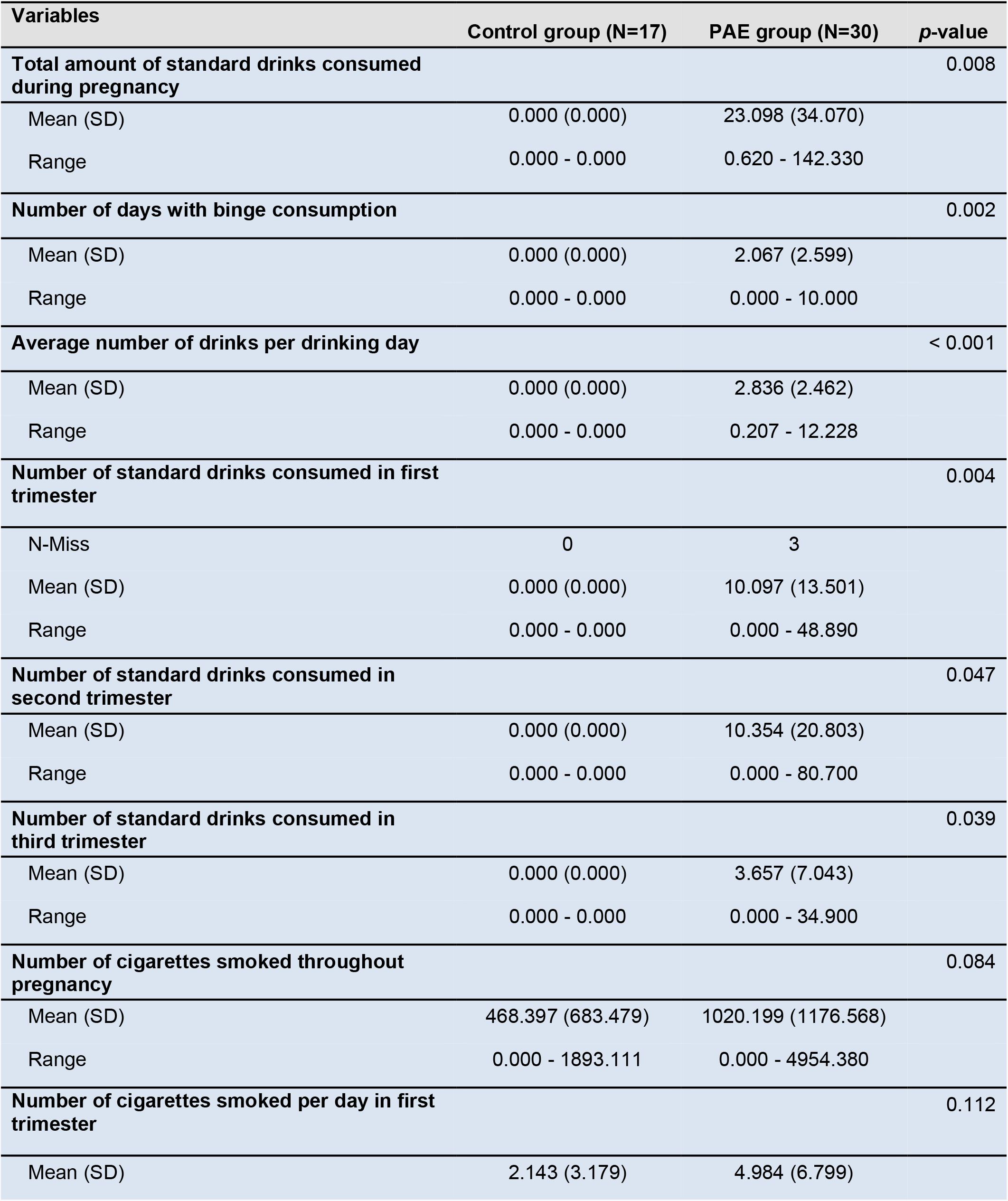
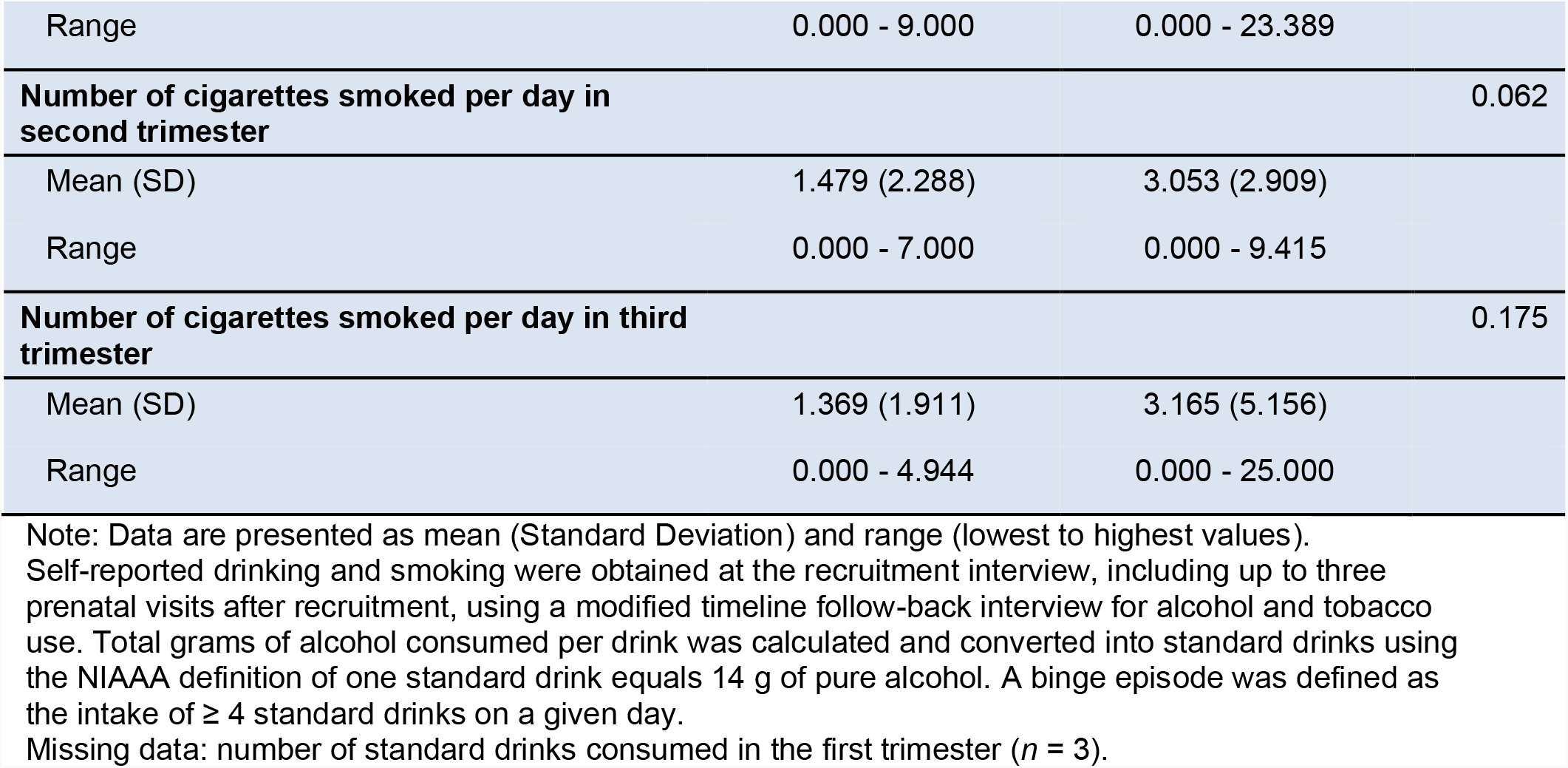
Quantity, frequency and timing of maternal alcohol and tobacco used

### Exposure

#### Cortical ROIs

Figure 1 shows the main effects of and interaction between PAE and PTE on cortical volume, thickness, and surface area across 68 bilateral ROIs.

**Fig. 1.**
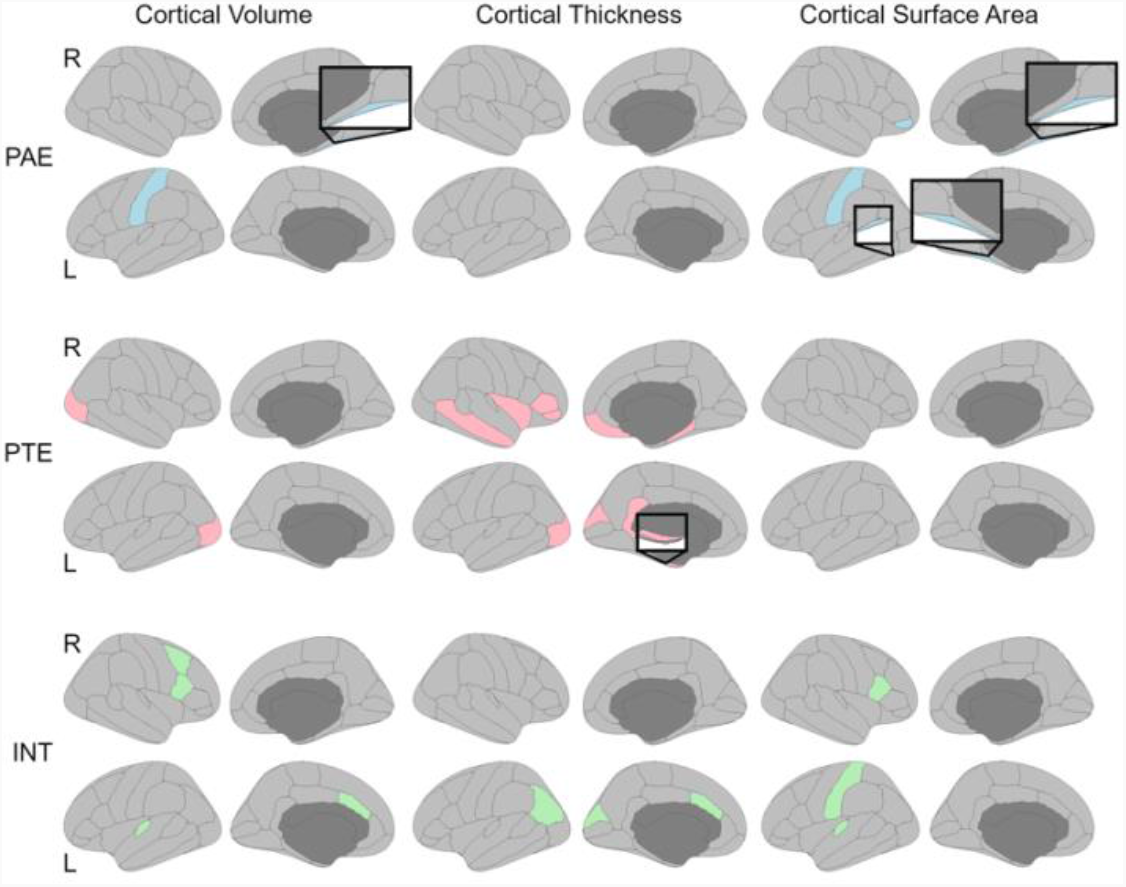
Cortical regions significantly associated with alcohol and tobacco exposure during pregnancy. L= left hemisphere. R= right hemisphere. PAE = prenatal alcohol exposure. PTE= prenatal tobacco exposure. INT = interaction.

PAE was associated with greater cortical volume in the left postcentral (p = 0.009) and right fusiform gyrus (p = 0.007). PAE was also associated with greater surface area in the left and right fusiform gyrus (p = 0.033 and p = 0.003, respectively), left postcentral gyrus (p = 0.043), and right pars orbitalis (p = 0.011).

PTE was associated with greater volume in the left and right lateral occipital cortex (p = 0.042 and p = 0.018, respectively). PTE was also associated with thicker cortices in right middle temporal (p = 0.043), right insula (p = 0.043), right pars triangularis (p = 0.011), right pars orbitalis (p = 0.014), right medial orbitofrontal (p = 0.030), right parahippocampal (p = 0.028), left lateral occipital (p = 0.040), left cuneus (p = 0.026), left isthmus cingulate (p = 0.033), and left entorhinal (p = 0.024) regions. PTE was also associated with greater surface area in the right temporal pole (p = 0.029).

Significant interactions of PAE with PTE in cortical volume were found in right caudal middle frontal (p = 0.028), right pars opercularis (p = 0.039), left transverse temporal (p = 0.029), and left caudal anterior cingulate (p = 0.048) regions. Interactions were also observed in cortical thickness for left inferior parietal (p = 0.049), left cuneus (p = 0.031), and left caudal anterior cingulate (p = 0.039). Interactions were observed in surface area for right pars opercularis (p = 0.027), left postcentral (p = 0.048), and left transverse temporal (p = 0.029) regions.

No main effects of PAE, PTE, or their interactions on cortical volume, thickness or surface area survived FDR correction.

#### Subcortical ROIs

PTE was associated with decreased volume in the left putamen (p = 0.039) and PAE × PTE interactions were found in the anterior, central, and posterior segments of the corpus callosum (p = 0.030, p = 0.039, p = 0.14), as well as the right cerebellum cortex (p = 0.031). However, no main effects of PAE, PTE, or their interactions on subcortical volume survived FDR correction. Figure 2 shows the main effects of PTE on cortical volume, thickness, and surface area across 68 bilateral ROIs.

**Fig. 2.**
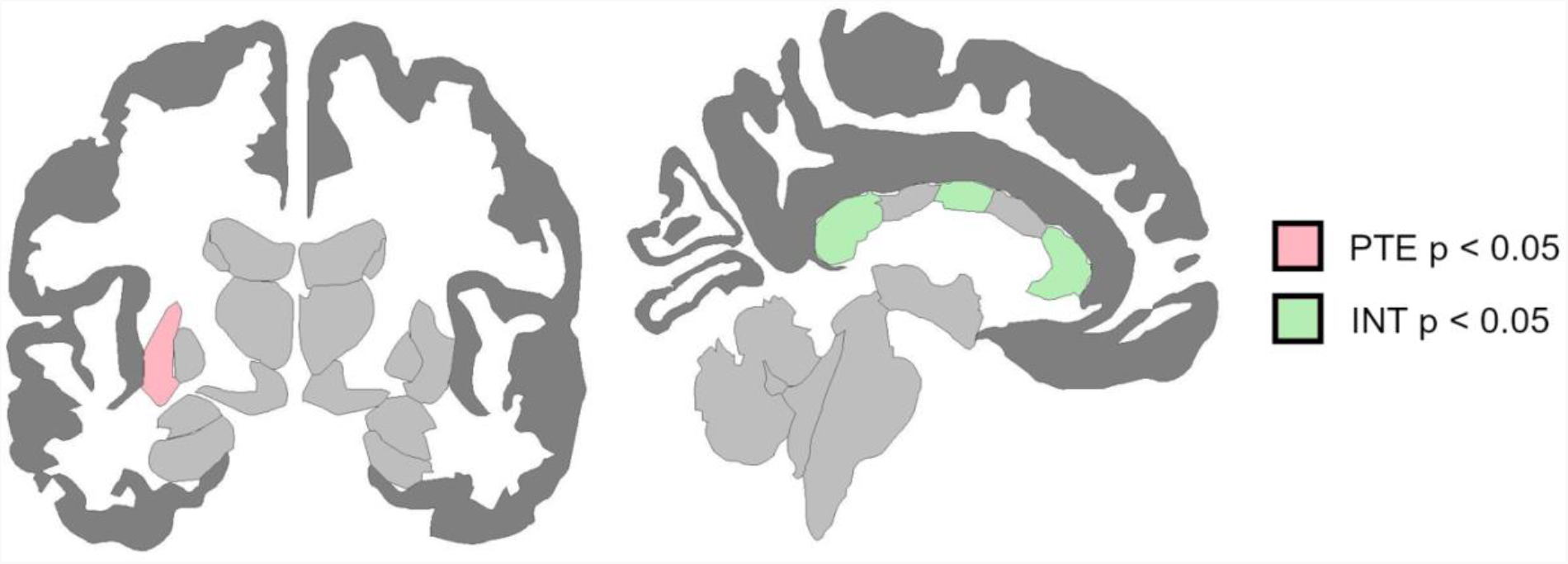
Subcortical regions significantly associated with tobacco exposure during pregnancy. PTE = prenatal tobacco exposure. INT = interaction.

**Fig. 3.**
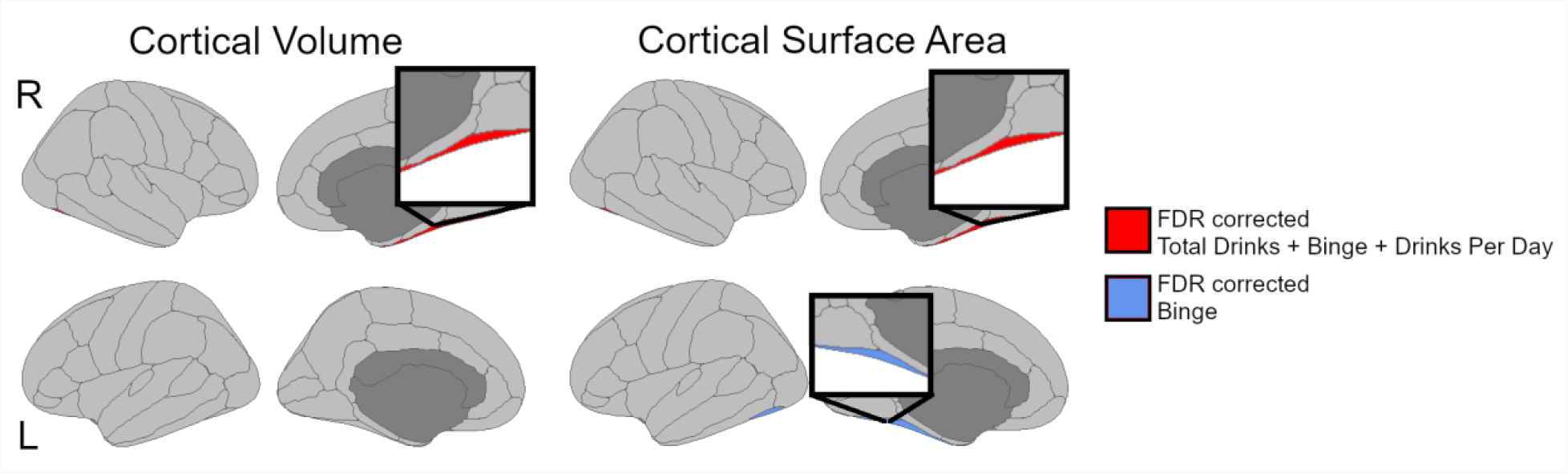
Cortical regions exhibiting significant associations with quantity and frequency of alcohol exposure during pregnancy (i.e., total amount of standard drinks consumed during pregnancy, binge drinking, the average number of drinks per drinking day).

**Fig. 4.**
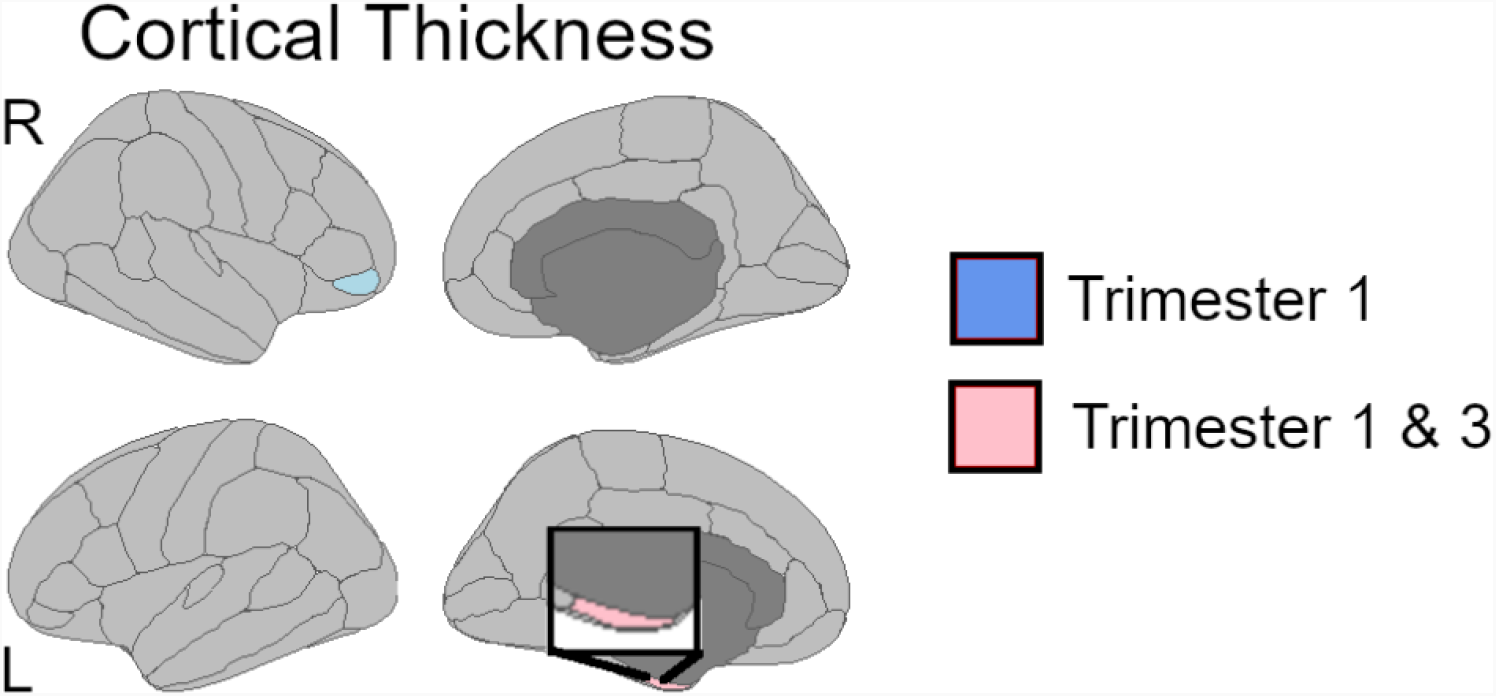
Cortical regions exhibiting significant associations with timing of tobacco exposure during pregnancy (i.e., trimester one and trimester 3 of pregnancy).

### Quantity, Frequency, Timing

#### Prenatal alcohol exposure

ROIs that showed significant group differences prior to FDR correction for either PAE main effect or the PAE × PTE interaction were studied further in relation to QFT of PAE. After FDR correction, the total amount of standard drinks consumed during pregnancy and the average number of drinks per drinking day were positively associated with cortical volume in the right fusiform gyrus (p = .003 and p = .010, respectively). Also, after FDR correction, binge drinking was positively associated with cortical volume in the right fusiform gyrus (p = .014). In addition, binge drinking was also positively associated with the surface area of bilateral fusiform gyrus (p = .040). The total amount of standard drinks consumed during pregnancy and the average number of drinks per drinking day were positively associated with surface area in the right fusiform gyrus (p = .028 and p = .010, respectively).

#### Prenatal tobacco exposure

ROIs that reached significance prior to FDR correction for either PTE main effect or the PTE × PAE interaction were further studied in relation to QFT of PTE. PTE in the first trimester was negatively associated with surface area of the right inferior parietal region (p = .029). PTE in the first and third trimester was negatively associated with cortical thickness in the right pars orbitalis (p = .010 and p = .027, respectively). PTE in the first trimester was negatively associated with volume of the right cerebellum cortex (p = .036). However, none of these associations survived FDR correction.

## Discussion

Several key findings emerge from this pilot study. Prior to correction for multiple testing, both PAE and PTE, as well as their interaction, were associated with a range of cortical and subcortical measures. The pattern of cortical and subcortical ROIs associated with PAE did not overlap with those associated with PTE. Furthermore, the profile of ROIs associated with the interactive effects of PAE with PTE exhibited a third profile different from PAE or PTE alone. These three ROI profiles suggest that the impact of PAE on brain structure at 6 years of age is unique from PTE, and their interactive effects are likely complex and not simply additive or synergistic. However, these findings remain preliminary, as none survived correction for multiple testing with the exception of increased right fusiform cortical volume and surface area with increased PAE quantity (binge and drinks per drinking day).

That these findings did not survive correction, suggests that our study, which included a range of alcohol exposures, was not sufficiently powered to demonstrate an association of exposures with brain measures. The possibility of PAE by PTE interaction deserves further study in larger samples, and further studies should also consider the higher PTE doses that naturally co-occur with PAE, compared to mono-substance use of PTE or PAE only.

It is noteworthy that despite the small sample size of this pilot study, the total amount of standard drinks consumed during pregnancy and the average number of drinks per drinking day, were positively associated with cortical volume in the right fusiform gyrus. This is in contrast to other neuroimaging studies, which have consistently demonstrated reduced cortical volume in children prenatally exposed to alcohol (Rajaprakash et al., 2014; Migliorini et al., 2015). Fusiform gyrus plays a role in high-level tasks related to visual processing, including processing of information about faces (Rangarajan et al., 2014; Weiner & Grill-Spector, 2012). Face processing is a critically important perceptual ability; identifying and interpreting facial emotions have a critical role in social functioning, and children with FASDs may have difficulty doing so (Lindinger et al., 2022). These findings, together with the observation of similar size effects for both PAE and PTE are novel, and deserve to be consolidated with more extensive work.

Several limitations deserve emphasis. Firstly, as described above, work on PAE and PTE that includes a range of different QFT features will require larger sample sizes to avoid false negative findings. Secondly, we did not include as potential moderators of our findings, a range of poverty-related factors, such as maternal exposure to interpersonal violence, childhood exposure to community violence, and food insecurity (Tomlinson et al., 2014; Stein et al., 2015; Koen et al., 2016; Donald et al., 2018; Gonzalez et al., 2020; Uban et al., 2020). Thirdly, given the focus on age 6, we are unable to determine whether the findings here are true null findings, or simply underpowered. Given the dynamic trajectories of brain volumes over time, with significant increases in cortical thickness at ages 5-6 years, and then subsequent thinning when approaching pubertal maturation, it is possible that the outcomes of PAE and PTE are also likely dynamic, and alterations in brain developmental trajectories following teratogen exposures may overlap. This may result in no observable group differences at certain ages, including around age 6, as in the case in this pilot study. Finally, exclusion criteria based on co-use of other substances other than alcohol and tobacco may have excluded women and their developing babies that may be most impacted by teratogens, and poly-use beyond the two most common ones (alcohol, tobacco) warrants more investigation to serve this likely high-risk sub-sample.

In conclusion, with regards to *exposure*, this pilot study was potentially insufficiently powered to demonstrate significant associations of community-levels of PAE, PTE, and their interaction with brain measures at age 6 years of age. However, it is noteworthy that uncorrected patterns of ROIs associated with PAE, PTE and their interaction comprised three unique profiles, suggesting that co-occurring exposures are likely highly complex and warrant greater investigation. The effect sizes for PAE and PTE were similar. Even in this small sample, significant positive associations of alcohol *quantity* with volume and surface area of the right fusiform were observed. Larger samples will be needed to fully delineate the impact of alcohol and tobacco exposure, as well as QFT characteristics on the developing brain, particularly when investigating naturally occurring patterns of PAE among the community, rather than solely focusing on very high levels of PAE found in clinical FASD studies.

## Data Availability

All data produced in the present work are contained in the manuscript.

## Acknowledgments

The authors gratefully acknowledge the contribution of the personnel and investigators of the Safe Passage Study. We would especially like to acknowledge the families for their participation in this study.

## Authors’ contributions

S.J. B. designed the study and participated in data collection. E.K., D.J., K.A.U., B.M. and S.A. assisted with data analysis and interpretation. K.A.U., D.J. and D.J.S. drafted the manuscript. L.B.K. assisted with formatting the manuscript. K.A.D., S.C.B., A.R., B.M., H.J.O. and E.R.S. all contributed to drafts of the article, with particular attention to areas of specific expertise, and all authors approved the final draft.

## Ethical standards

Ethical approval for human subject research was obtained from the Human Research Ethics Committee of the Faculty of Health Sciences of Stellenbosch University (REF 248/2014). Informed consent and assent were obtained from parents/guardians and participants before enrolment.

